# Mitochondrial DNA Copy Number Is Associated With Visual Field Progression in Primary Open-Angle Glaucoma

**DOI:** 10.64898/2026.09.05.26362328

**Authors:** Anh H. Pham, Katelyn M. Kane, Douglas R. da Costa, Felipe A. Medeiros

## Abstract

**Purpose:** To investigate the association between mitochondrial DNA (mtDNA) copy number (CN) and the rate of visual field progression in patients with primary open-angle glaucoma.

**Design:** Retrospective cohort study

**Participants:** 200 patients with primary open-angle glaucoma (POAG) with a mean of 11 years of longitudinal clinical data and DNA extracted from peripheral blood.

**Methods:** DNA extraction was performed on peripheral blood leukocytes collected from 200 POAG patients who had an average follow-up of 11 years. Mitochondrial CN was quantified by multiplex quantitative TaqMan PCR using custom primers and probes against cytochrome *b* and ß-2 microglobulin as measures of mitochondrial and nuclear genes, respectively. Univariable and multivariable linear regression models were used to evaluate the association between mtDNA CN and rates of visual field progression, quantified by the slope of standard automated perimetry (SAP) mean deviation (MD) over time.

**Main Outcome Measures:** Association of mtDNA CN with MD slope.

**Results:** Mean mtDNA CN was 107.6 copies per cell (interquartile range, 70.5–126.7). Higher ln(mtDNA CN) was associated with slower rates of MD loss in both univariable (*β* = 0.29, p < 0.001) and multivariable analyses (*β* = 0.25, p = 0.003). Higher peak IOP was independently associated with faster visual field progression in both analyses. A significant interaction was observed between mtDNA CN and peak IOP (*β* = −0.054, p < 0.001), with the association between lower mtDNA CN and faster SAP progression being more pronounced at lower levels of peak IOP.

**Conclusion:** Lower mtDNA CN was associated with faster SAP progression, with the association being most pronounced at lower levels of peak IOP. These findings support mtDNA CN as a potential systemic biomarker of mitochondrial genomic reserve and susceptibility to visual field loss.

## 1 Introduction

Glaucoma is a progressive neurodegeneration of the optic nerve characterized by marked loss of retinal ganglion cells (RGC). Primary open-angle glaucoma (POAG) is the most common subtype, affecting over 68 million people worldwide, and predominates in Western countries. ^1^ While older age, nuclear genetic variants, epigenetic aging ^2^, sub-Saharan African ancestry, family history, and high myopia are all risk factors for POAG, ^3^ intraocular pressure (IOP) is the only modifiable risk factor that is targeted by medical, laser, and surgical therapy. Despite effective reductions in IOP, prospective longitudinal studies estimate that 33-66% ^4–6^ of patients will have persistent visual field loss and 5-10% of patients will progress to legal blindness in their lifetime. ^7,8^ These studies highlight the importance of exploring IOP-independent mechanisms involved in glaucoma progression. Although past studies have identified blood pressure ^9–11^, ocular perfusion pressure ^12^, and high polygenic risk scores ^13,14^ as systemic risk factors for progression, few cellular markers of progression risk have been identified.

Impaired mitochondrial function has emerged as a common feature of POAG. ^15–18^ Mitochondria are dynamic multifunctional organelles that supply cells with bioenergetic fuel through oxidative phosphorylation (OXPHOS) and fatty acid metabolism. Other important cellular pathways regulated by mitochondria include programmed cell death, oxidative stress, viral defense, and the stimulator of interferon genes (STING) inflammatory cascade. The mitochondrial DNA (mtDNA) exists as multiple copies (hundreds to thousands) within the cell and encodes protein subunits for 4 of the 5 OXPHOS complexes. Pathogenic mutations and large deletions in the mitochondrial genome ^17,19,20^ and mutations in nuclear genes encoding mitochondrial proteins ^21^ have been identified in POAG patients, implicating mitochondrial deficiency in RGC loss. Extensive literature supports the loss of mtDNA copy number (CN) in aging, neurodegenerative diseases, and cognitive decline ^22–25^ suggesting that mtDNA CN is a dynamic biomarker of mitochondrial fitness and cellular health.

Mitochondrial genetics and function influence visual outcomes across several ocular diseases. A higher mtDNA CN is protective from vision loss in families with Leber’s Hereditary Optic Neuropathy ^26^ whereas low CN is more prevalent in patients with severe dry age-related macular degeneration. ^27^ Because mtDNA CN can be assayed from peripheral blood leukocytes, it offers an accessible systemic marker for mitochondrial reserve that can complement routine clinical tests. In glaucoma, lower oxidative respiration in peripheral blood leukocytes also correlates with faster visual field loss. ^16^ Thus, we investigated the association between mtDNA CN and rates of visual field progression using multivariable linear regression models adjusted for relevant demographic and clinical covariates.

## 2 Methods

### 2.1 Study group

This study involved a cohort of patients who were part of the Duke Glaucoma Registry, a large database of electronic medical records developed at Duke University, Durham, North Carolina. The database consists of adults 18 years of age or older with a glaucoma or glaucoma-suspect diagnosis who were evaluated at the Duke Eye Center or its satellite clinics between January 2009 and June 2023. The database has been described in detail elsewhere and has been used for several studies investigating risk factors for glaucoma. ^3,18–21^ A subset of participants was recruited for prospective blood sample collection during regular clinic visits. Blood samples were collected, processed, and stored for subsequent analyses, with DNA extraction and molecular analyses performed at the John P. Hussman Institute for Human Genomics (HIHG) at the University of Miami, Miami, Florida. The Duke University and University of Miami Institutional Review Boards approved the study. Informed consent was obtained from all participants. All methods adhered to the tenets of the Declaration of Helsinki for research involving human subjects and were conducted in accordance with regulations of the Health Insurance Portability and Accountability Act of 1996.

The database used for this study contained clinical information from baseline and follow-up visits, including patient diagnostic and procedure codes, medical history, smoking history, best-corrected visual acuity, slit-lamp biomicroscopy, IOP measurement using Goldmann applanation tonometry (Haag-Streit), central corneal thickness (CCT), gonioscopy, ophthalmoscopy examination, optic disc photographs, and the results of all standard automated perimetry (SAP) and OCT examinations. Standard automated perimetry testing was performed with the Humphrey Field Analyzer (Carl Zeiss Meditec) using the Swedish Interactive Thresholding Algorithm 24-2 fast strategy. Reliable SAP results were defined as having fixation loss rate of less than 25% and false-positive rate of < 15%. Visual fields were reviewed manually for artifacts such as lid and rim artifacts, fatigue effects, inappropriate fixation, and evidence that the visual field results were the result of a disease other than glaucoma.

### 2.2 Participant Selection

This study included a subset of participants from the Duke Glaucoma Registry who were recruited for prospective blood sample collection. Patients were contacted during their regular clinic visits and were invited to provide blood samples for genetic analysis. Selection criteria included a diagnosis of primary open-angle glaucoma (POAG) in both eyes, confirmed by International Classification of Diseases codes at the baseline visit, as well as the availability of longitudinal data in the Duke Glaucoma Registry. A total of 10 ml of blood was collected from each participant, processed, and stored for future analysis. Blood collections took place between June 2021 and June 2023.

### 2.3 DNA Extraction and Quality Control

Immediately after collection, whole blood samples were centrifuged to separate the different components. The separated components, including plasma, buffy coat, and red blood cells, were then frozen at −80°C for preservation. At a later stage, the buffy coat was thawed and processed for DNA extraction after centrifugation at 1500g for 10 minutes to concentrate the cells. Genomic DNA was extracted using the QIAamp DNA Blood Mini Kit (Qiagen) following the manufacturer’s protocol. DNA concentrations were quantified on a Qubit 2.0 Fluorometer (Life Technologies) using the Qubit dsDNA HS Assay Kit (Life Technologies). A subset of DNA samples was evaluated qualitatively by agarose gel electrophoresis to confirm high molecular weight bands suitable for analysis. Sample concentration was normalized to 50 ng/ml and arrayed in Azenta 0.5 ml barcoded tubes in racks of 96 by HIHG. For the qPCR assay, the DNA concentration and quality was confirmed on NanoDrop spectrophotometer (Thermo Fisher Scientific) and diluted to 10ng/ul with DNase- and RNase-free water.

### 2.4 TaqMan Quantitative Real-Time PCR and mtDNA CN Calculation

Quantitative real-time PCR (qPCR) was performed using a multiplex TaqMan assay to quantify cytochrome *b (*cyt *b)* and ß-2 microglobulin (ß2M) as measures of mtDNA and a single-copy nuclear gene (nDNA), respectively. The qPCR assay was performed using the QuantStudio 5 Real-Time PCR System (Applied Biosystems) with the following cycling conditions: 5 min at 95°C and 40 cycles of 95°C for 15 seconds, 60°C for 30 seconds, and 72°C for 30 seconds. The 20 *μ*L reaction mixture contained 10 ng of DNA, 1X SsoAdvanced Universal Probe multiplex supermix (Bio-Rad), 1X ROX Reference Dye (Invitrogen), 250 nM primers, and 200 nM TaqMan probes. A plasmid containing known CN of cytochrome *b* and ß-2 microglobulin was included in each plate to construct a standard curve. For the mitochondrial gene, primers in the D-loop were avoided because of its high error rate and sequence heterogeneity. ^28^ Forward primers included 5’-GCCTATATTACGGATCATTTCTCTACT-3’ (cyt *b)* and 5’-CACGTC ATCCAGCAGAGAATGGAAAGTC-3’ (ß2M). Reverse primers included 5’-GCCTATGAAGGCTGTTGCTATAGT-3’ (cyt *b)* and 5’-CAATTCTCTCTCCATTCTTCAGTAAGTCAAC-3’ (ß2M). PrimeTime TaqMan probes were designed using specified dyes (FAM emission 520 or SUN™ emission 554) with a double-quencher system ZEN™ and Iowa Black fluorescence quencher (IBFQ) from Integrated DNA Technologies (IDT). TaqMan probes included 5’-SUN™ / ZEN™ / CCTGAAACATCGGCATTATCCTCCTGCT / IBFQ-3’ (cyt *b) and* 5’-FAM / ZEN™ / ATGTGTCTGGGT TTCATCCATCCGACA / IBFQ-3’ (ß2M). The mtDNA:nDNA ratio was calculated and multiplied by 2 (due to the diploid nature of nDNA) to obtain CN per cell. Assay was performed in technical triplicate and CN was averaged across replicates ensuring a standard deviation of ≤15% across replicates.

### 2.5 Statistical Analysis

Descriptive statistics were used to summarize demographic and clinical characteristics of the cohort. Continuous variables are presented as mean ± standard deviation (SD) and categorical variables as counts and percentages. The primary outcome was the rate of visual field progression, defined as the slope of change in standard automated perimetry (SAP) mean deviation (MD) over time (dB/year), estimated using ordinary least squares regression. For analysis, parameters from the eye with the fastest progression during follow-up were used. The primary exposure variable was mitochondrial DNA copy number (mtDNA CN) per cell measured from buffy coat samples. Because mtDNA CN exhibited a right-skewed distribution, values were natural log-transformed prior to modeling.

Associations between natural log-transformed mtDNA CN and MD slope were first evaluated in univariable linear regression models and subsequently in multivariable models adjusting for potential confounders selected a priori, including age at blood collection (per 10-year increment), sex, race, follow-up duration, baseline MD, peak intraocular pressure (IOP), and central corneal thickness (CCT). Eye-specific variables corresponded to measurements from the eye with the fastest progression.

To examine whether the association between mtDNA CN and progression differed according to IOP levels, an interaction term between ln(mtDNA CN) and peak IOP was introduced into the multivariable model. Continuous variables involved in the interaction were mean-centered to facilitate interpretation of main effects. Statistical significance of interaction effects was assessed using joint Wald tests. Model performance was evaluated using the coefficient of determination (R^2^). All statistical analyses were performed using Stata.

## 3 Results

A total of 200 patients were included in the study. **Table 1** shows demographic and baseline characteristics for the cohort. Eyes with the fastest progression showed a rate of mean deviation (MD) loss of −0.59 ± 0.93 (mean ± SD) decibel (dB) annually on SAP while the slowest eyes exhibited average loss of −0.12 ± 0.25 dB per year. A more negative slope in dB/year indicates faster progression. The average age at blood collection was 72.6 years ± 9.3, with a mean follow-up time of 11 years ± 6.5. The study population was racially diverse, with 21.5% of patients self-identifying as Black, 2.5% as Asian, and the remainder as White.

**Table 1:** Demographic and Clinical Characteristics of Cohort.

| Variable | Mean (SD) | Median | IQR |
| --- | --- | --- | --- |
| *Demographics* |  |  |  |
| Age at collection, years | 72.6 (9.3) | 73.6 | 67.7-79.5 |
| Follow-up, years | 11.0 (6.5) | 10.4 | 6.0-15.3 |
| Female sex, n (%) | 117 (58.5%) | – | – |
| Race, n (%) |  |  |  |
| White | 152 (76.0%) | – | – |
| Black | 43 (21.5%) | – | – |
| Asian | 5 (2.5%) | – | – |
| *Mitochondrial DNA* |  |  |  |
| Average mtDNA per cell | 107.6 (63.7) | 95.5 | 70.5-126.7 |
| $\ln(\text{mtDNA per cell})$ | 4.47 (0.79) | 4.56 | 4.26-4.84 |
| *Visual Field Progression (dB/year)* |  |  |  |
| Fastest eye MD slope | -0.59 (0.93) | -0.34 | -0.69 to -0.10 |
| Slowest eye MD slope | -0.12 (0.25) | -0.08 | -0.20 to -0.01 |
| *Baseline Visual Field* |  |  |  |
| Fastest eye baseline MD, dB | -5.4 (6.7) | -3.2 | -7.3 to -0.9 |
| Slowest eye baseline MD, dB | -4.4 (6.4) | -2.3 | -5.7 to -0.5 |
| *Intraocular Pressure* |  |  |  |
| Fastest eye peak IOP, mmHg | 22.7 (8.9) | 20.0 | 17.0-25.5 |
| Slowest eye peak IOP, mmHg | 21.5 (6.9) | 20.0 | 17.0-24.0 |
| *Central Corneal Thickness* |  |  |  |
| Fastest eye CCT, $\mu\text{m}$ | 545.2 (37.1) | 546.3 | 525.3-566.5 |
| Slowest eye CCT, $\mu\text{m}$ | 547.4 (41.0) | 546.2 | 524.0-572.0 |

Two hundred patients were included in the study. Negative slope indicates worsening visual field loss. Data are presented as mean ± standard deviation (SD) unless otherwise indicated. MD = mean deviation; IOP = intraocular pressure; CCT = central corneal thickness; IQR = interquartile range.

The distribution of measured mtDNA CN for all study participants is shown in **Figure 1**. Mean mtDNA CN was 107.6 ±63.7 copies per cell, with most values concentrated between approximately 50 and 150 copies per cell. These values are consistent with those reported in previous studies using blood-derived samples, in which mtDNA CN estimates vary according to sample composition and, in particular, the degree of platelet contamination. ^29,30^

**Figure 1:**
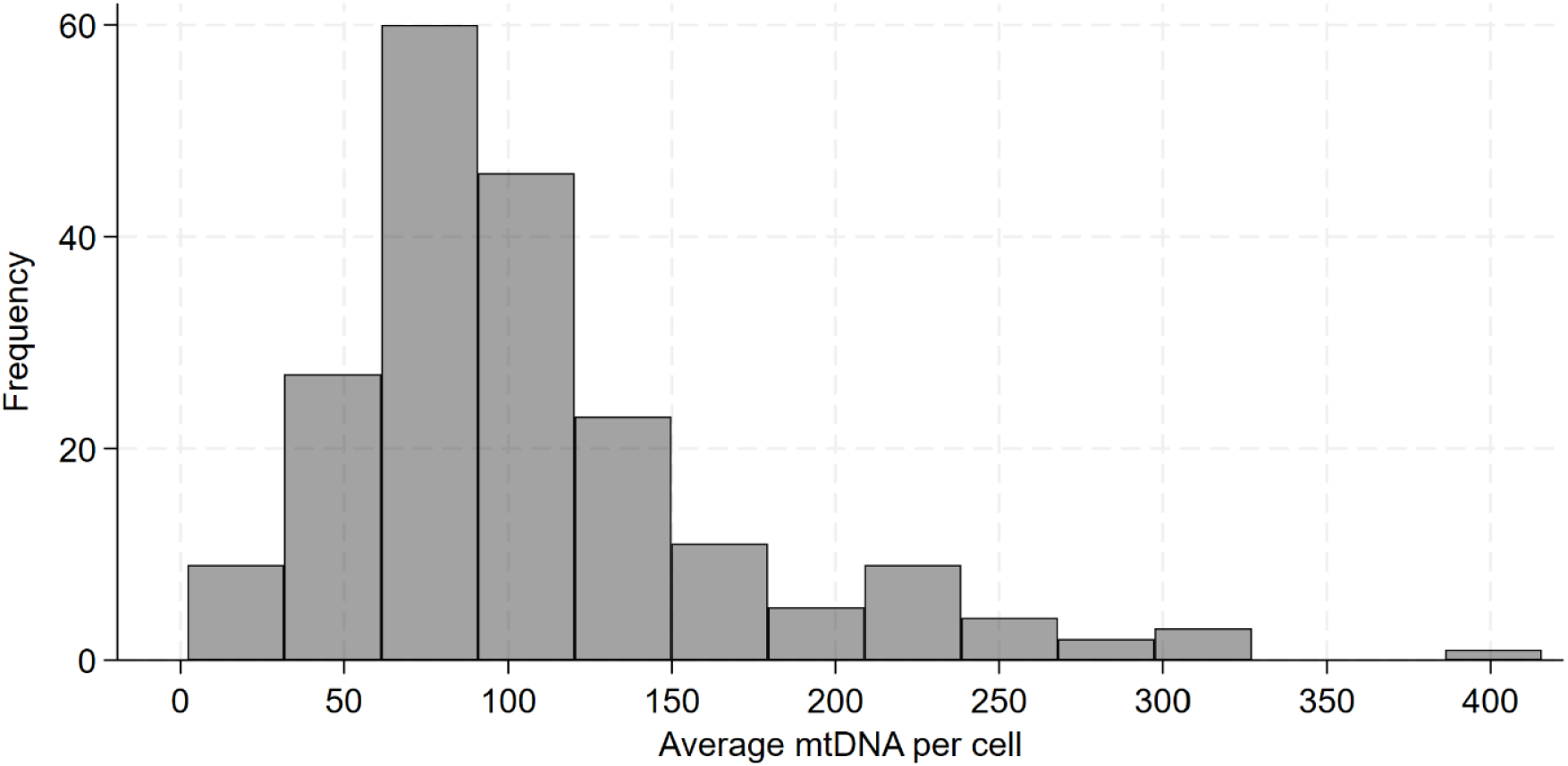
Frequency Distribution of Mitochondrial Genome in Study Population. Histogram of mtDNA CN per cell across 200 study participants. The x-axis represents the mtDNA CN per cell and the y-axis shows the number of participants with that copy number.

The relationship between mtDNA CN and the rate of visual field progression is illustrated in **Figure 2**. Lower mtDNA CN was associated with more negative MD slopes, corresponding to faster rates of SAP progression. To formally evaluate this association, we fitted univariable and multivariable linear regression models with MD slope as the outcome (**Table 2**). The multivariable model adjusted for relevant demographic and clinical covariates, including follow-up duration and ocular characteristics from the eye with the fastest rate of visual field progression. Because the distribution of mtDNA CN was right-skewed, mtDNA CN was natural log-transformed before regression modeling.

**Table 2:** Results of Linear Regression Models for the Association Between Mitochondrial DNA and Fast Glaucoma Progression.

| Variable | Univariable |  | Multivariable |  |
| --- | --- | --- | --- | --- |
|  | Coefficient | p-value | Coefficient | p-value |
| ln(mtDNA per cell) | 0.2914 | <0.001 | 0.2475 | 0.003† |
| Age at collection, per 10 years | -0.0311 | 0.661 | -0.073 | 0.300 |
| Follow-up time, per year | 0.0230 | 0.024 | 0.0186 | 0.066 |
| Sex (Female) | -0.0506 | 0.706 | -0.0626 | 0.636 |
| Race (White) | -0.1592 | 0.302 | 0.1453 | 0.339 |
| Baseline MD, dB | 0.0224 | 0.022 | 0.0156 | 0.108 |
| Peak IOP, mmHg | -0.0156 | 0.035 | -0.0220 | 0.003† |
| CCT, $\mu\text{m}$ | 0.0035 | 0.052 | 0.0024 | 0.175 |
| ln(mtDNA) x Peak IOP | – | – | -0.0542 | <0.001 |

Results from univariable and multivariable linear regression analyses examining factors associated with visual field progression rate (mean deviation slope in dB/year). Positive coefficients indicate slower progression. Variables were mean-centered prior to creating interaction terms to ensure interpretable main effects. †Joint Wald tests were performed for variables involved in the interaction term: for ln(mtDNA per cell), $F(2, 190) = 12.10$ , $p < 0.0001$ ; for Peak IOP, $F(2, 190) = 12.07$ , $p < 0.0001$ . $R^2 = 0.202$ , $p < 0.001$ .

**Figure 2:**
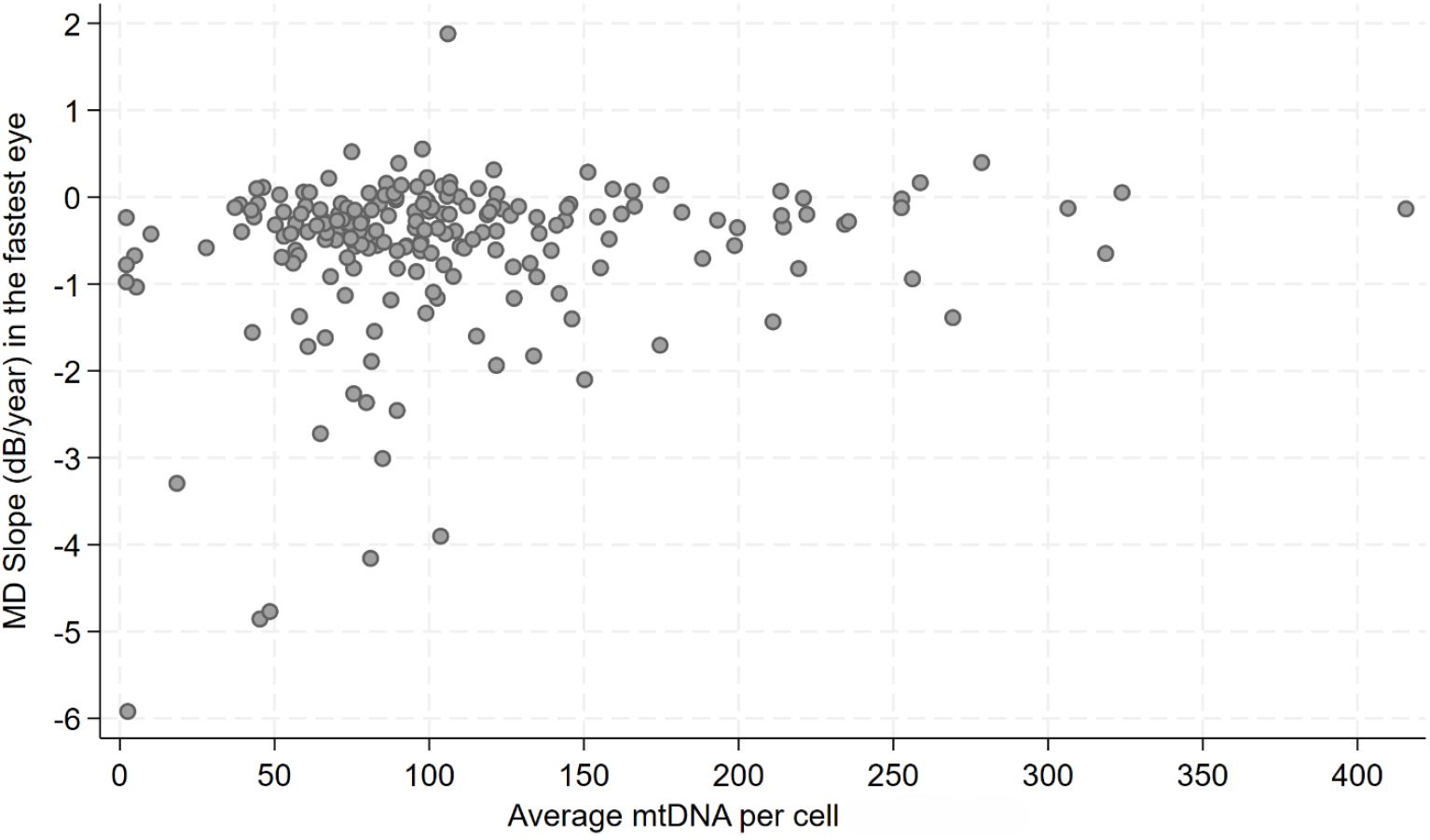
Scatter Plot of Mean Deviation Slope versus Average Mitochondrial DNA CN. Scatter plot shows the relationship between average mtDNA CN per cell (x-axis) and mean deviation slope (y-axis) across all study participants. Each point represents an individual participant.

Higher ln(mtDNA CN) was significantly associated with a less negative MD slope in both univariable (*β* = 0.2914, p < 0.001) and multivariable analyses (*β* = 0.2475, p = 0.003), indicating that greater mtDNA CN was associated with slower visual field progression after adjustment for potential confounders. Higher peak IOP was independently associated with a more negative MD slope in both univariable (*β* = −0.0156, p = 0.035) and multivariable analyses (*β* = −0.0220, p = 0.003). Follow-up duration and baseline MD were significantly associated with MD slope in univariable analyses but were no longer statistically significant after multivariable adjustment. Age, sex, race, and central corneal thickness were not significantly associated with MD slope in either model.

Predicted rates of visual field progression across levels of mtDNA CN, derived from the multivariable regression model, are shown in **Figure 3**. Lower mtDNA CN was associated with progressively more negative predicted MD slopes. At the covariate values used for model prediction, an mtDNA CN of 75 copies per cell corresponded to a predicted MD slope of approximately −0.61 dB/year, compared with approximately −0.44 dB/year at 150 copies per cell. Thus, a twofold higher mtDNA CN within the range commonly observed in the study population was associated with an approximately 0.17 dB/year, or 28%, slower predicted rate of visual field loss.

**Figure 3:**
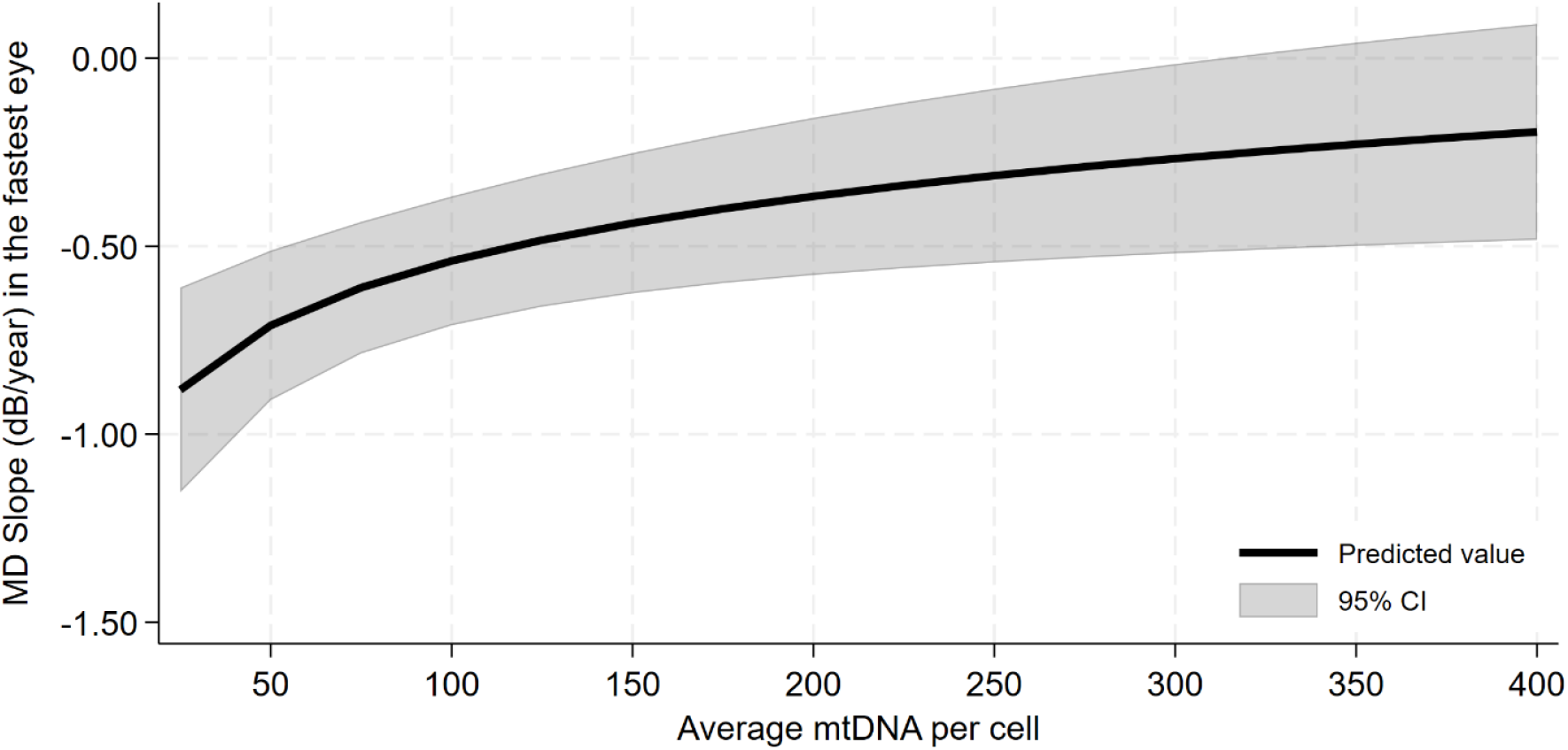
Predicted Visual Field Progression by Mitochondrial DNA Level. Predicted rates of mean deviation slope based on average mtDNA levels. Graph was computed from the multivariable linear regression model. The dark line represents the predicted mean deviation slope value and the shaded area represents the 95% confidence interval. The model is adjusted for sex, race, peak intraocular pressure, baseline mean deviation, central corneal thickness, and follow-up time until blood collection. Model predictions are based on the following mean covariate values: age = 73 years, follow-up = 11 years, sex = Female, race = White, baseline mean deviation = −5.4 dB, peak intraocular pressure = 22.7 mmHg, and central corneal thickness = 545 *μ*m.

We next examined whether the association between mtDNA CN and visual field progression varied according to peak IOP by including an interaction term between ln(mtDNA CN) and peak IOP in the multivariable model (**Table 2**). The interaction was highly significant (p < 0.001), indicating that the relationship between mtDNA CN and MD slope differed substantially across levels of peak IOP. As illustrated in **Figure 4**, the association was strongest at lower IOP levels and became progressively attenuated with increasing peak IOP. An increase in mtDNA CN from 75 to 150 copies per cell corresponded to an approximately 0.46 dB/year difference in predicted MD slope at a peak IOP of 15 mmHg, 0.27 dB/year at 20 mmHg, and only 0.09 dB/year at 25 mmHg. Thus, the magnitude of the association was approximately 41% smaller at 20 mmHg and 80% smaller at 25 mmHg compared with that observed at 15 mmHg. These findings suggest that the association between mitochondrial genomic reserve and visual field progression may be particularly relevant in patients whose disease progresses despite relatively low IOP.

**Figure 4:**
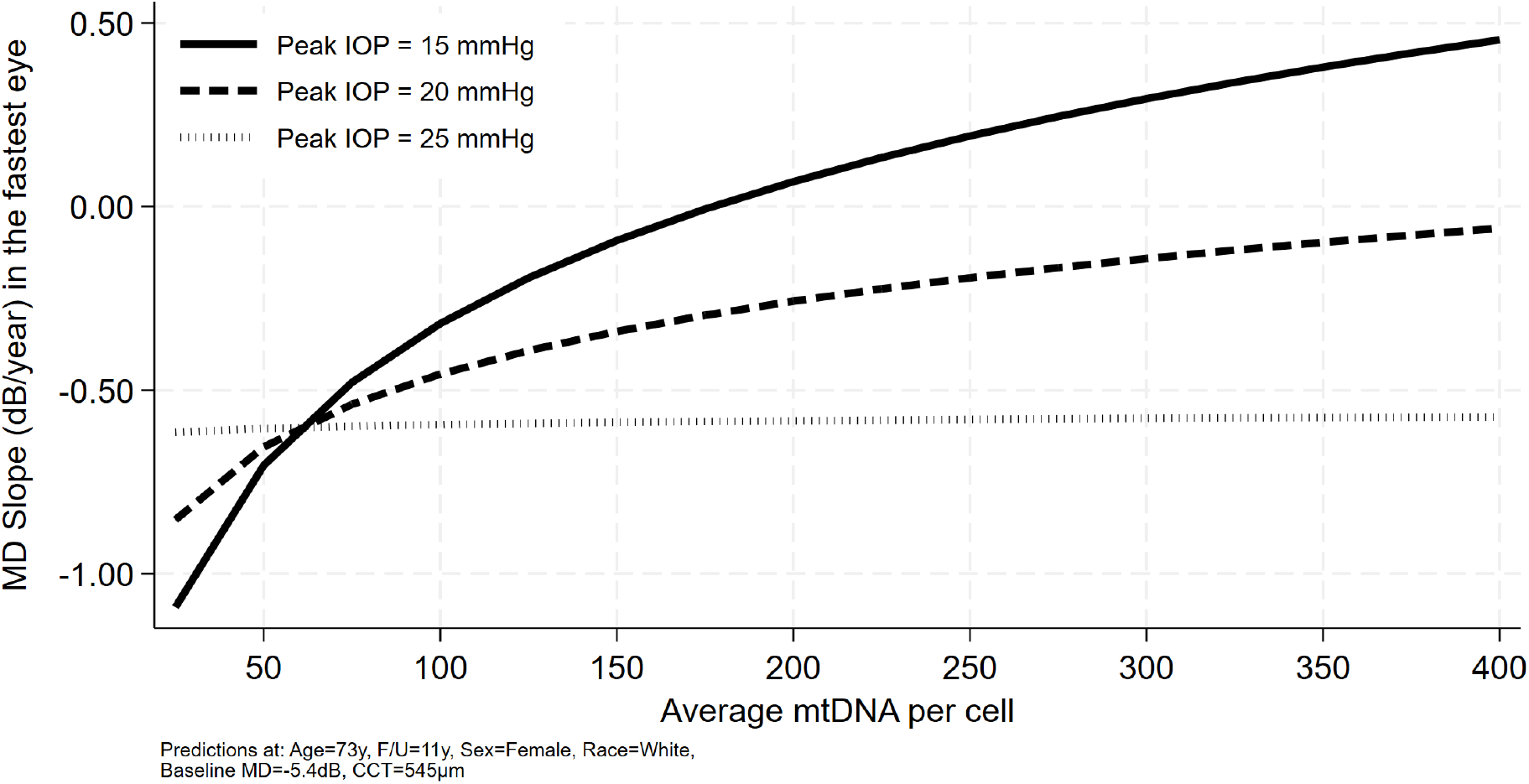
Predicted Visual Field Progression by Mitochondrial DNA Level is Dependent on Peak IOP. Predicted visual field progression in relation to mtDNA CN per cell, stratified by peak IOP. Predicted rate of MD loss annually (dB/year) is plotted against different levels of mtDNA (x-axis) for three peak IOP values. Graph is derived from the multivariable linear regression model with interaction term. Predictions use mean covariate values for age, sex, follow-up, CCT, race, and baseline MD.

## 4 Discussion

This study demonstrates a significant association between lower mtDNA CN and faster visual field progression in patients with POAG followed for a mean of 11 years. Lower mtDNA CN was associated with more negative MD slopes in both univariable and multivariable analyses, and the association persisted after adjustment for relevant demographic and clinical covariates. Peak IOP was also independently associated with faster progression. Importantly, we identified a significant interaction between mtDNA CN and peak IOP, with the association between lower mtDNA CN and faster visual field loss being substantially stronger at lower IOP levels. Together, these findings support mtDNA CN as a potential systemic biomarker of susceptibility to glaucomatous visual field progression, particularly when IOP-related risk is relatively low.

Our findings are consistent with a growing body of evidence implicating systemic mitochondrial dysfunction in glaucoma. Petriti et al. ^16^ demonstrated that reduced mitochondrial respiratory capacity in peripheral blood leukocytes was associated with more rapid glaucomatous visual field loss. Other studies have reported impaired complex I activity and ATP synthesis in lymphoblasts from patients with POAG. ^31,32^ Inoue-Yanagimachi and colleagues also reported an association between the blood mtDNA/nDNA ratio and ocular blood-flow measures in a subgroup of patients with severe open-angle glaucoma, further supporting a relationship between systemic mitochondrial abnormalities and ocular phenotypes. ^33^ Importantly, that association was observed in a subgroup rather than across the entire open-angle glaucoma cohort and therefore should be interpreted cautiously.

The study most directly comparable to ours is that of Vallbona-Garcia et al. ^29^, who reported lower mtDNA CN in buffy-coat DNA from patients with high-tension glaucoma compared with control participants. Their findings provide independent evidence that reduced peripheral-blood mtDNA CN may characterize a subset of patients with glaucoma. Our study extends these observations by examining mtDNA CN in relation to longitudinal rates of visual field loss rather than glaucoma status alone. Several methodological differences may also be relevant when comparing the studies, including differences in study populations and mtDNA quantification. Vallbona-Garcia et al. ^29^ quantified mtDNA using the mitochondrial D-loop, whereas our assay avoided the D-loop because of its greater sequence heterogeneity and susceptibility to error in mtDNA CN estimation. ^28^ In addition, the longitudinal follow-up in our cohort allowed estimation of individual MD slopes over many years, providing a direct assessment of the relationship between mtDNA CN and subsequent rates of functional loss.

An important implication of our findings is that mtDNA CN may provide information about susceptibility to visual field progression beyond routinely measured clinical characteristics. The association between mtDNA CN and MD slope remained significant in the multivariable model, while higher peak IOP was independently associated with faster visual field loss. The magnitude of the association with mtDNA CN was also potentially clinically meaningful. At the covariate values used for model prediction, an mtDNA CN of 75 copies per cell corresponded to a predicted MD slope of approximately −0.61 dB/year, compared with approximately −0.44 dB/year at 150 copies per cell, representing an approximately 28% slower predicted rate of visual field loss at the higher mtDNA CN level (**Figure 3**). These findings raise the possibility that mtDNA CN could complement established clinical measures in identifying patients at greater risk of rapid functional deterioration.

The most notable finding in this study was the significant interaction between mtDNA CN and peak IOP. The association between lower mtDNA CN and faster visual field progression was greatest at lower levels of peak IOP and became progressively attenuated as peak IOP increased (**Figure 4**). For example, increasing mtDNA CN from 75 to 150 copies per cell corresponded to an approximately 0.46 dB/year difference in predicted MD slope at a peak IOP of 15 mmHg and 0.27 dB/year at 20 mmHg, but only 0.09 dB/year at 25 mmHg. Thus, the magnitude of the mtDNA-associated difference was approximately 80% smaller at a peak IOP of 25 mmHg than at 15 mmHg. This interaction suggests that mitochondrial reserve may be particularly relevant to disease progression when the contribution of elevated IOP is relatively limited.

This observation is biologically consistent with emerging evidence that mitochondrial dysfunction may contribute substantially to glaucomatous neurodegeneration at lower IOP levels. Petriti et al. ^16^ further reported that respiratory impairment in these cells was particularly pronounced among patients with normal-tension glaucoma (NTG). Our findings extend these observations by demonstrating that a readily measurable marker of mitochondrial genomic reserve is differentially associated with visual field loss according to IOP level. Although these results do not establish that mitochondrial dysfunction is the predominant mechanism of progression in NTG, they support the concept that the relative contribution of mitochondrial vulnerability may vary across the spectrum of IOP-related disease. If confirmed in independent longitudinal cohorts, mtDNA CN could therefore help identify a biologically distinct subgroup of patients in whom IOP-independent mechanisms contribute substantially to progression.

Multiple genetic studies have implicated impaired mitochondrial homeostasis in NTG. Optineurin (OPTN) and TANK-binding kinase 1 (TBK1) are nuclear-encoded genes that regulate the clearance of damaged mitochondria (mitophagy) and maintain mitochondrial quality control. Heritable mutations in these genes account for 1% to 2% of NTG cases. ^34^ Similarly, mutations in OPA1, a nuclear gene critical for mitochondrial inner membrane fusion and cristae junction stability, are associated with a 2-fold higher risk of NTG. ^35,36^ Within the mitochondrial genome, point mutations in complex I subunits and large-scale deletions are enriched in POAG and NTG patients. ^20,37^ Because these genes regulate mitophagy, mitochondrial dynamics, and genomic integrity, these findings collectively implicate disruption in the mitochondrial life cycle as a mechanism of mtDNA depletion and accelerated RGC loss. Future studies are required to identify which of these pathways (impaired biogenesis, aberrant mitophagy, or abnormal membrane dynamics) contributes to glaucoma pathogenesis and progression.

Therapeutic strategies that target mitochondrial function or the mitochondrial genome may merit consideration as adjunctive treatment when additional IOP reduction offers limited benefit. Both mtDNA CN and respiratory capacity may be modifiable risk factors. Experimental models have shown that mitochondrial biogenesis and metabolic activity respond to exercise ^38^, caloric restriction ^39^, and pharmacological agents that activate the peroxisome proliferator-activated receptor *γ* coactivator 1*α* (PGC-1*α*), including metformin ^40^, nicotinamide precursors ^41^, and resveratrol. ^42^ Nicotinamide supplementation, in particular, has shown promising effects on mitochondrial mass, respiratory capacity, and axonal transport ^41^, and is currently being evaluated in several global clinical trials in glaucoma. ^43^ Our data predict that the benefit would be greatest in patients with low mtDNA CN whose disease progresses despite relatively low peak IOP, but results from these ongoing trials will be needed to determine whether improving mitochondrial health slows progression.

Our study has several limitations, some of which we have discussed previously. ^2^ Because blood was collected after the observed visual field progression, we cannot establish causality, as low mtDNA CN may have preceded the progression or developed as a consequence of disease trajectory. In addition, because most patients with POAG were under treatment, we cannot exclude an effect of treatment on the mitochondrial genome. The cohort was drawn from a single center, so the findings may not generalize to other populations; analyses in larger, multicenter cohorts will be necessary. Although the buffy coat was carefully isolated, residual platelets and minor red-blood-cell contamination may have influenced mtDNA CN estimates. Buffy-coat mtDNA CN was also quantified at a single time point and may not reflect mitochondrial content in RGC; studies in ocular tissue will be needed to determine whether blood mtDNA CN is a valid surrogate for RGC metabolic status. Finally, systemic comorbidities, medications, and smoking history were not incorporated into our models and may represent confounders.

In conclusion, lower mtDNA CN was independently associated with faster visual field progression in patients with POAG, with the association being most pronounced at lower peak IOP. Measurement of mtDNA CN in buffy-coat may therefore provide a readily accessible systemic marker of mitochondrial genomic reserve and a potential biomarker of susceptibility to glaucoma progression. Further validation in independent longitudinal cohorts will be important to establish its prognostic utility.

## Data Availability

All data produced in the present study are available upon reasonable request to the authors

